# Analysis of Intervention Effectiveness Using Early Outbreak Transmission Dynamics to Guide Future Pandemic Management and Decision-Making in Kuwait

**DOI:** 10.1101/2021.01.07.21249409

**Authors:** Michael G. Tyshenko, Tamer Oraby, Joseph Longenecker, Harri Vainio, Janvier Gasana, Walid Q. Alali, Mohammad AlSeaidan, Susie Elsaadany, Mustafa Al-Zoughool

## Abstract

Severe Acute Respiratory Syndrome Coronavirus 2 (SARS-CoV-2) is a World Health Organization designated pandemic that can result in severe symptoms and death that disproportionately affects older patients or those with comorbidities. Kuwait reported its first imported cases of COVID-19 on February 24, 2020. Analysis of data from the first three months of community transmission of the COVID-19 outbreak in Kuwait can provide important guidance for decision-making when dealing with future SARS-CoV-2 epidemic wave management. The analysis of intervention scenarios can help to evaluate the possible impacts of various outbreak control measures going forward which aim to reduce the effective reproduction number during the initial outbreak wave. Herein we use a modified susceptible-exposed-asymptomatic-infectious-removed (SEAIR) transmission model to estimate the outbreak dynamics of SARS-CoV-2 transmission in Kuwait. We fit case data from the first 96 days in the model to estimate the basic reproduction number and used Google mobility data to refine community contact matrices. The SEAIR modelled scenarios allow for the analysis of various interventions to determine their effectiveness. The model can help inform future pandemic wave management, not only in Kuwait but for other countries as well.

## 1. Introduction

The Kuwait Ministry of Health reported the country’s first four, imported COVID-19 cases on February 24, 2020. Contact tracing showed all were travel-related cases from Iran. Two of the cases were asymptomatic when tested (KUNA, 2020). In the initial stages of the domestic outbreak contact tracing as well as home and institutional quarantine measures were used to limit viral transmission from travelers entering the country (Al-Shammari et al., 2020). After just three weeks the confirmed number of cases increased to 112, and after six weeks a total of 556 cases were identified including the first death from COVID-19 on April 4, 2020 (Virusncov.com, 2020). Kuwait, like other countries dealt with repatriation, bringing more than 50,000 Kuwaiti citizens from around the world back by May 7, 2020.

Due to the rapidly increasing number of detected cases and undetected community transmission occurring within the first few weeks of the outbreak, Kuwait government officials acted quickly to implement several non-pharmaceutical interventions (NPIs) to reduce person-to-person transmission. Control measures to contain the community spread of SARS-CoV-2 included: closure of a wide range of institutions (schools, universities, government offices and non-essential businesses), full border lockdown, partial curfew, and lockdown.

Despite early and aggressive control measures community transmission continued to occur. By July 31, 2020 the Ministry of Health reported a total of 66,957 cases and 447 deaths (Worldometer, 2020). Kuwait and other countries faced with a lack of case data and early testing results implemented NPIs during a time of high uncertainty.

To improve decision-making and better understand the effectiveness of NPIs we used social contact rates within a deterministic model fitted to early case data. To realize this objective we developed a susceptible (S), exposed but not infectious (E), infected but asymptomatic (A), infected and symptomatic (I), and removed (R) due to recovery, isolation, hospitalization, or death (SEAIR) model. The social contact rates were improved by using Google mobility data for various interventions including school closures, business closures, lockdown and curfew (Google, 2020). The analysis can improve decision-making for NPIs during future pandemic waves.

## 2. Model and Methods

### 2.1. Model Description

We established a SEAIR disease transmission model to depict the spread of SARS-CoV-2 in Kuwait. We fit case data of the first 96 days of the epidemic to a multinomial likelihood function dependent on the SEAIR model to estimate the relevant model’s parameters. We also derive and estimate the effective basic reproduction number which is very close to the effective basic reproduction number since the fraction of susceptible in the beginning of the epidemic stays close to one in this epidemic and the former also includes the fraction who are not practicing social isolation. We used it to understand what would have been the course of the epidemic under various NPI scenarios. We also used it to find the likely time of the peak of the epidemic if it was not for the 20-days lockdown.

The deterministic model is constructed from a set of ordinary differential equations (ODE) with the five states or compartments (S, E, A, I, R). (Figure 1).

**Figure 1.**
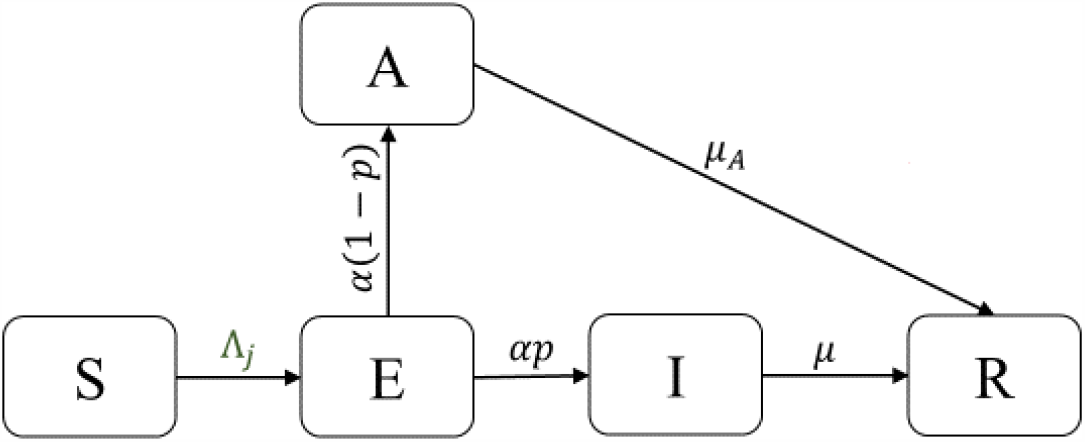
Schematic illustration of the SEAIR model of COVID-19. S is the compartment of susceptible individuals. E is the compartment of exposed and latent individuals but not infectious yet. A is the compartment of asymptomatic individuals. I is the infected individuals who are still able to interact with people (mildly infected) and choose not to isolate themselves. R is the removed individuals due to recovery, hospitalization, isolation, or death.

All compartments are classified by age. The force of infection *Λ*_*j*_ is given by equation (6). The rates α, µ_*A*_, and µ are the rates of removal from the E, A, and I, respectively. The fraction *p* is the probability of becoming symptomatic and infectious.

Compartments were split into four age groups: children (0–20 years), denoted by a (*c*) subscript, young adults (21–55 years) denoted by a (*ya*) subscript, adults (56–65 years) denoted by an (*a*) subscript, and seniors (66 years and older), denoted by a (*s*) subscript.

for *j* = *c, ya, a, s*; where the force of infection inflected on a susceptible individual in the *j*th age group is given by

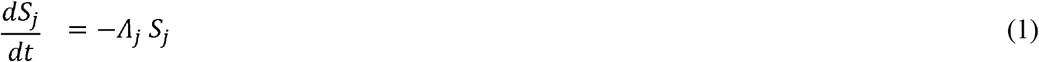

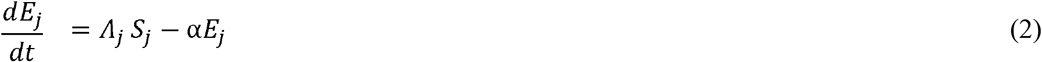

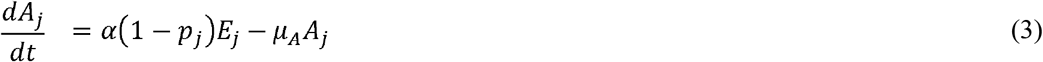

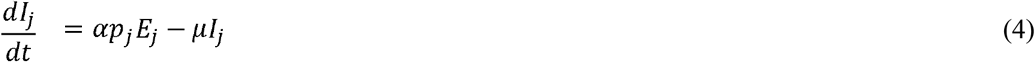

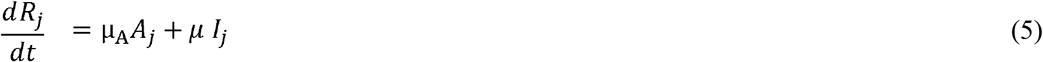

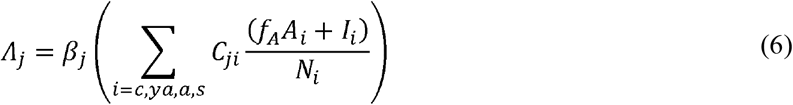

for *j* = *c, ya, a, s*.

The force of infection is dependent on the contact rates that would be affected by the changes in contact patterns due to social distance and lockdowns. We use here a function by replacing *β*_*j*_ in equation (6) by *β*_*j*_ *ρ*(*t*) where *ρ*(*t*) is the fraction of those who are not practicing social isolation.

We used the Next-generation matrix method (Diekmann et al., 1990) to derive *R*_0_ for the ODE model. The basic reproduction number is given by R_0_ = ***ρ***(***FV***^−1^), where

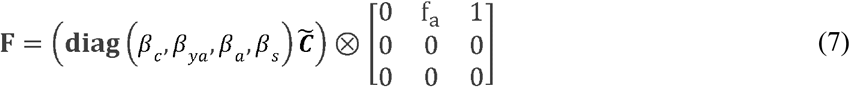

and

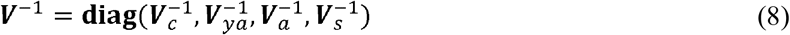

is a block diagonal matrix whose diagonal block matrices are given by

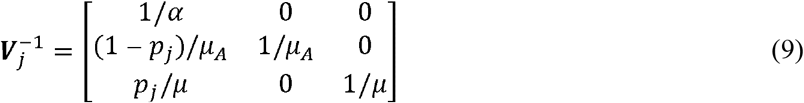

for *j = c, ya, a, s*, with (⨂) being the Kronecker product. The matrix 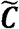 is the effective contact matrix with the change in mobility.

### 2.2. Model’s Parameters Estimation

We estimate the probability of successful transmissions β_*j*_ for *J = c, ya, a, s*; along with both the parameters *f*_*A*_ and µ. We also estimated the initial data of the exposed, asymptomatic and infectious individuals in each age group. Let the vector of parameters be denoted by Θ. We use values from the literature for the parameters α = 1/5 (Guo et al., 2020), and *µ*_*A*_ = 1/14 (Zhou et al., 2020). We use the results of Davies et al., (2020) to initialize the probability of transmission and the fraction of those becoming symptomatic p_*j*_ (clinical fraction). Google mobility report data was used to estimate the fraction of those who are not practicing social distance ρ(*t*) (Google, 2020).

We use a multinomial likelihood function

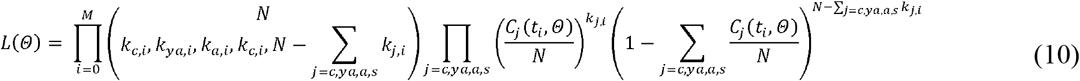

where *C*_*j*_ (*t*_*i*_, *Θ*) is the predicted number of cases by the SEAIR model (1-6) and N is the population size. We found the maximum-likelihood estimator of the parameters *β*_*c*_, *β*_*ya*_, *β*_*a*_, *β*_*s*_, *f*_*A*_ and µ. While there is no guarantee that the optimal solution is no more than arguments for a local maximum value of the likelihood function, we used various procedures to make sure what we are finding is the best possible. Estimating the parameters was performed on the negative log-likelihood function using different methods provided by the Global Optimization Toolbox in MATLAB (MathWorks, Inc., USA). In one of the trials, we used a Latin Hyper-Cube sampling to find a good initial point to start the optimization algorithms via the multi-start procedure that, in its turn, starts the search from that point as well as other 99 initial points generated by the procedure. We also used the Genetic Algorithm in the same toolbox and ran it parallel on 12 CPU cores with the appropriate options as provided by the documentation of the function in MATLAB. In the Genetic Algorithm part, we commanded a mixed-integer optimization for the initial data. We used the bootstrap method to quantify uncertainty in the estimates and calculate confidence intervals (Efron and Tibshirani, 1993).

### 2.3. Kuwait Data Description

We used available case data from the beginning of the COVID-19 epidemic in Kuwait on February 24, 2020 up to the beginning of the lockdown (May 10, 2020), a period of 96 days to inform NPI effectiveness. During this first period of the epidemic (until day 76) NPIs and policies implemented tended to be homogenous (ie. NPIs were applied uniformly to the entire population). Testing had similar turnouts and reporting rates were very close giving confidence in the data collected for fitting within our deterministic model. Country case data was provided by the Ministry of Health, Government of Kuwait (Government of Kuwait, 2020).

## 3. Results

The parameters used in the following simulations are based on fitting the model’s parameters that are relevant to the COVID-19 epidemic in the first 96 days starting on February 24, 2020, see Figure 2 and Table 1. We estimated that it is highly likely that the epidemic was initiated by one exposed child and one exposed adult, in addition to one infected young adult and one infected senior.

**Table 1.** The SEAIR model’s parameters, values/estimates, standard errors or ranges and their sources.

| Parameters | Values/Estimates | SE/Ranges | Sources |
| --- | --- | --- | --- |
| $(\beta_c, \beta_{ya}, \beta_a, \beta_s)$ | (0.0846, 0.1081, 0.2659, 0.5392) | (0.0241, 0.0154, 0.0476, 0.1126) | Estimated |
| $\mu$ | 0.8685 | 0.1797 | Estimated |
| $f_A$ | 0.3854 | 0.1095 | Estimated |
| $(p_c, p_{ya}, p_a, p_s)$ | (0.25, 0.50, 0.56, 0.66) | Ranges (0.12 – 0.38, 0.18 – 0.60, 0.49 – 0.76, 0.57 – 0.82) | Calculated based on estimates in Davies et al. (2020) |
| $\alpha$ | 1/5 | Range (1/7, 1/3) | Gou et al., 2020 |
| $\mu_A$ | 1/14 | Range (1/37, 1/14) | Zhou et al., 2020 |

**Figure 2.**
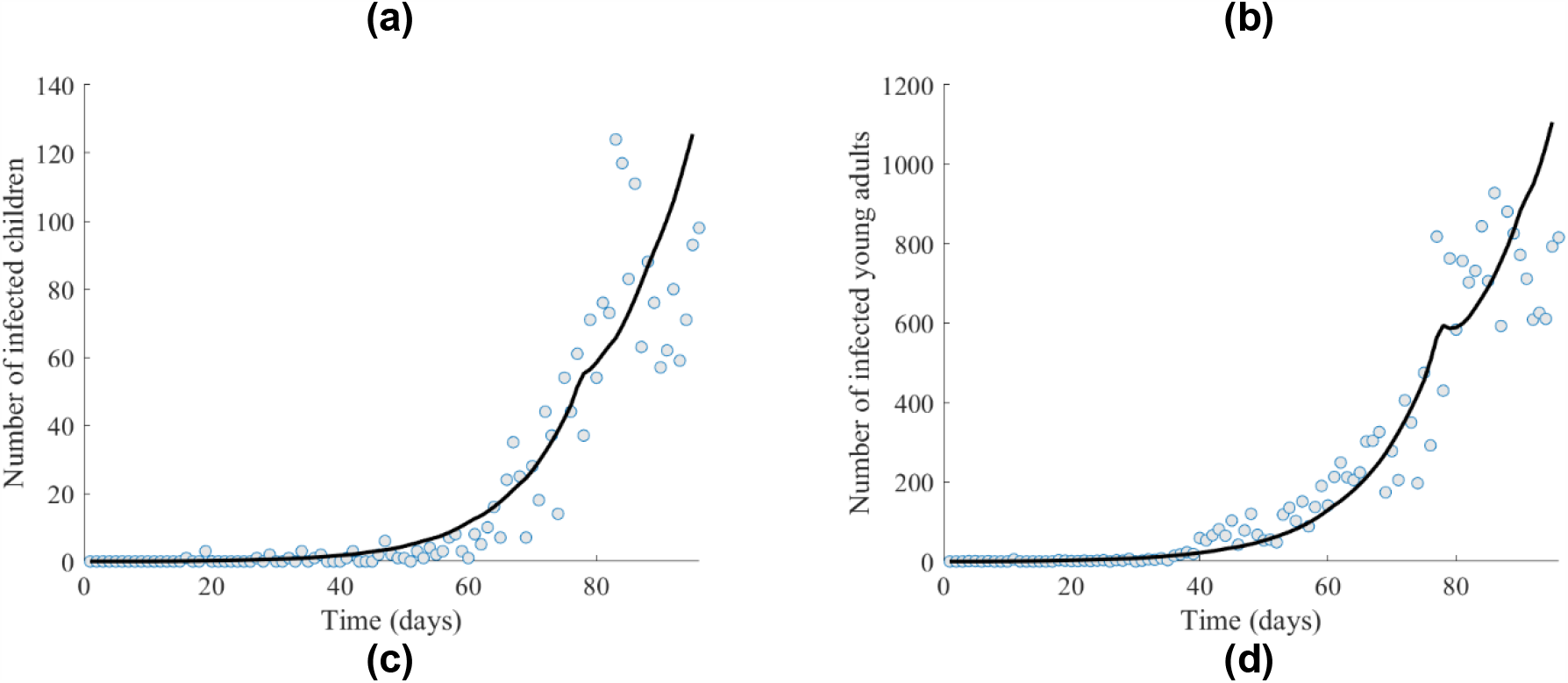
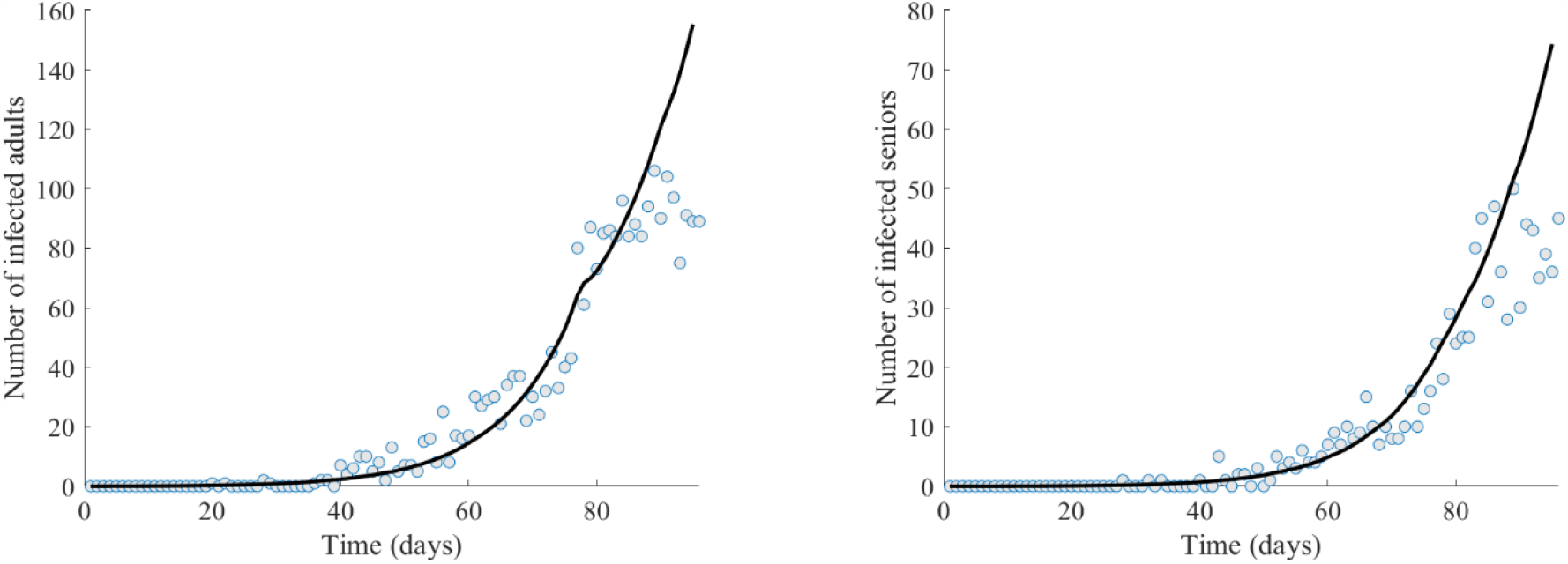
Fitted curves for the four age groups (a) children, (b) young adults, (c) adults, and (d) seniors.

The following figure shows the best fit curves as a solution of the SEAIR model (1-6) against the actual reported cases (Figure 2).

Based on the estimated parameters, the basic reproduction number started at *R*_0_ = 11.86 in the beginning of the epidemic and reached *R*_0_ = 4.69 at the end of the lockdown (Figure 3a). We also projected the epidemic with the measures and social isolation levels before the lockdown forward from day 76 to find the likely time of the peak of the epidemic. We used also the bootstrap sampled data to make a boxplot of its time (Figure 3b). We found the mean of the peak’s time to be July 3^rd^ and the median to be July 11^st^. In overall, it ranges from June 26^th^ to August 5^th^.

**Figure 3.**
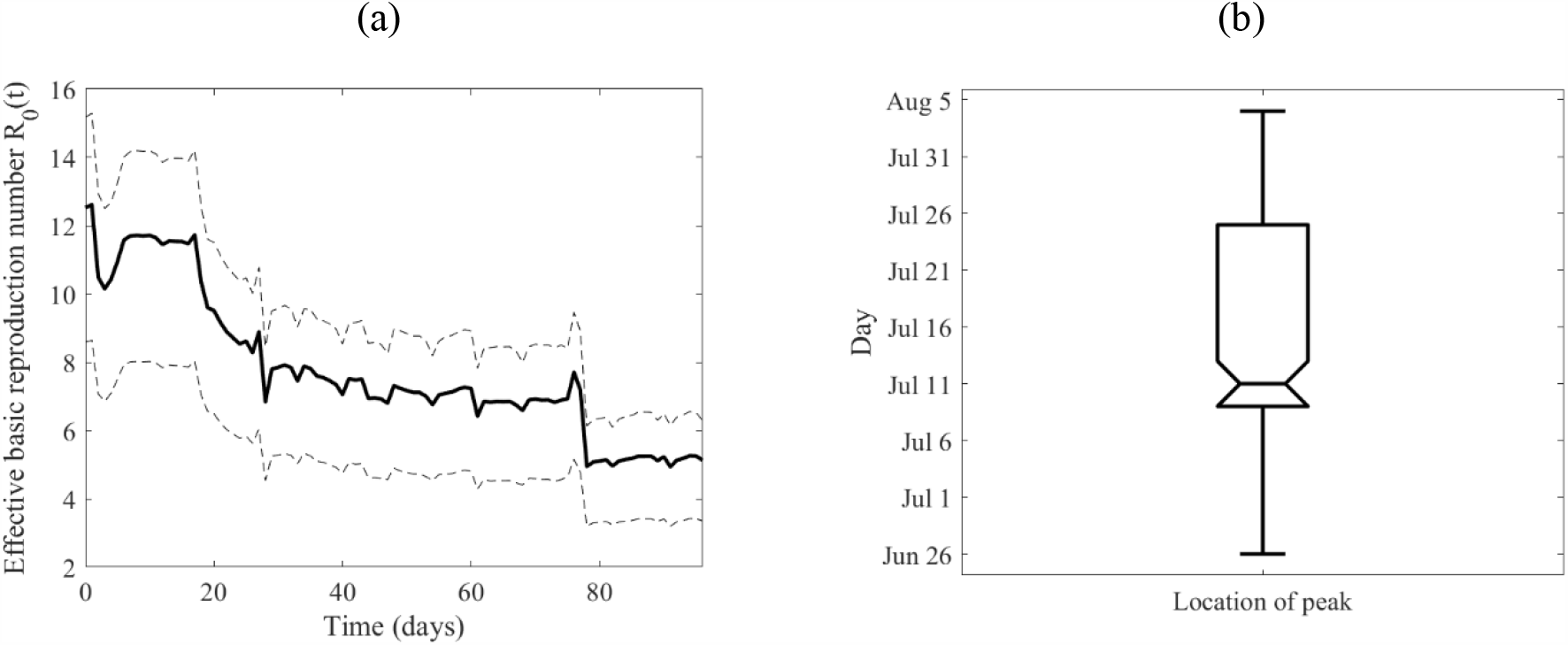
(a) The estimated effective basic reproduction number *R* (*t*) over the first 97 days in Kuwait with a 95% confidence interval. (b) A boxplot of the peak time if there were no lockdown and the measures before the lockdown were continued.

### 3.1. Timeline and Scenarios

Social contact rates used in the SEAIR model for Kuwait were taken from our previously derived social contact matrix (Al-Zoughool et al., 2020; Prem et al., 2019) and the effects on social contact rates were updated using Google mobility data. Table 2 and Figure 4 show a timeline of the various measures (interventions and transmission control strategies) implemented by the government of Kuwait during the first wave of COVID-19. Modifications to population-level, social contact rates were informed by using Google mobility data for Kuwait (Google, 2020). We estimated the effects of the measures on social contact rates and applied these as percentage changes to all age groups.

**Table 2.**
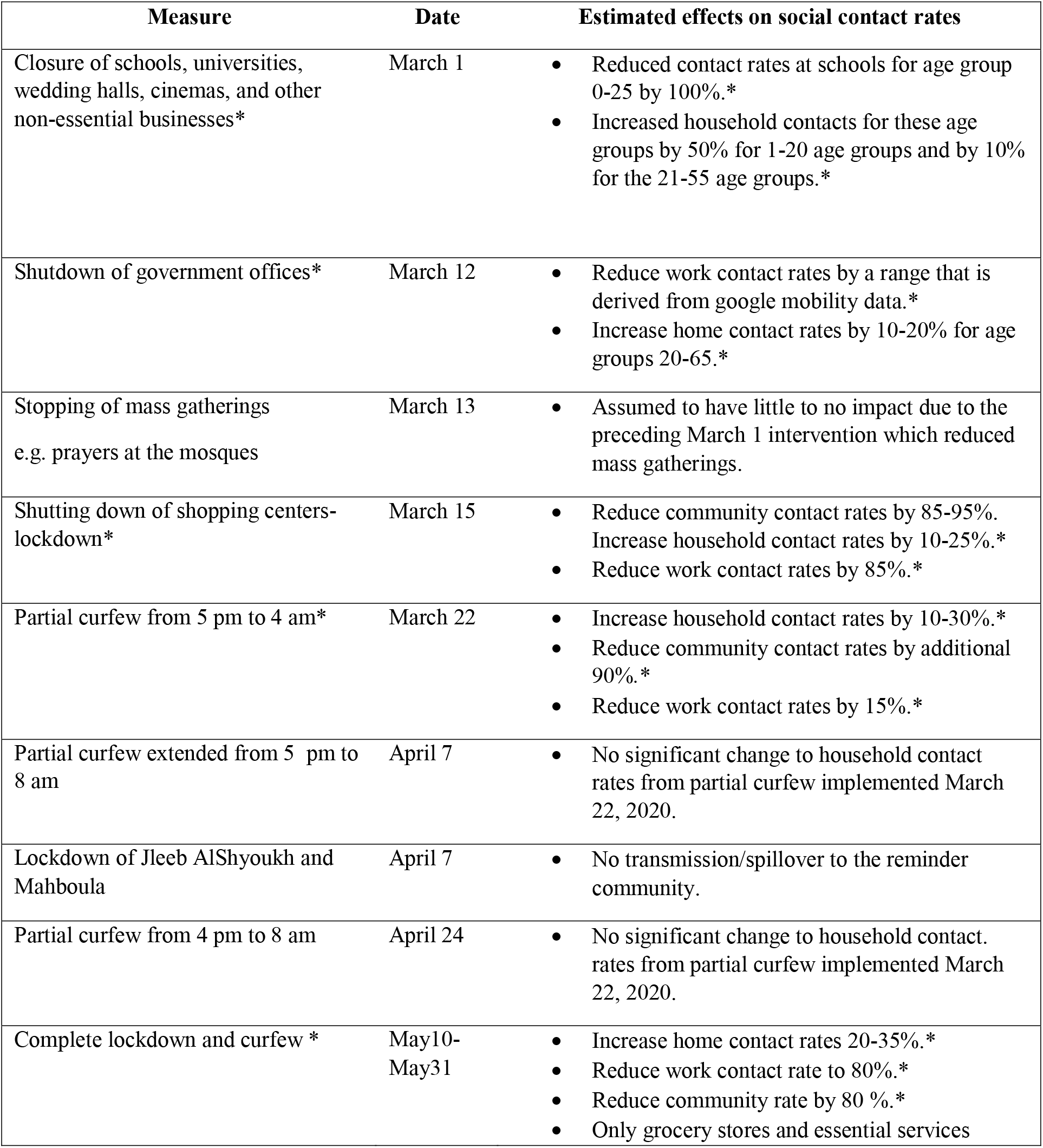
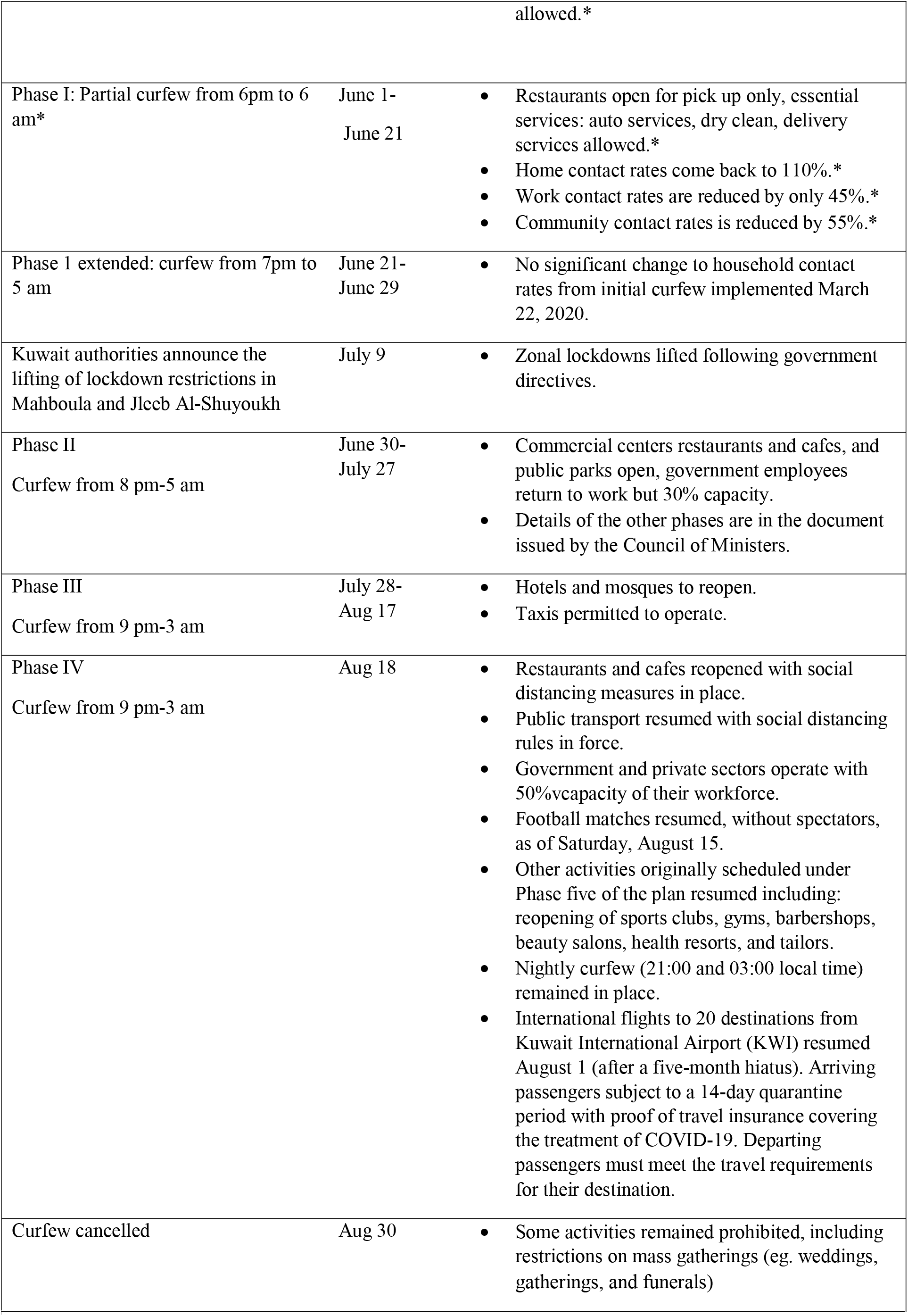
Timeline of measures implemented from March 1 to August 30 2020 to reduce COVID-19 transmission in Kuwait and the estimated effects on social contact rates (as percentage change) informed by Google mobility data. Major changes to social contact (*) were noted for several measures and their associated estimated effects using Google mobility data.

**Figure 4.**
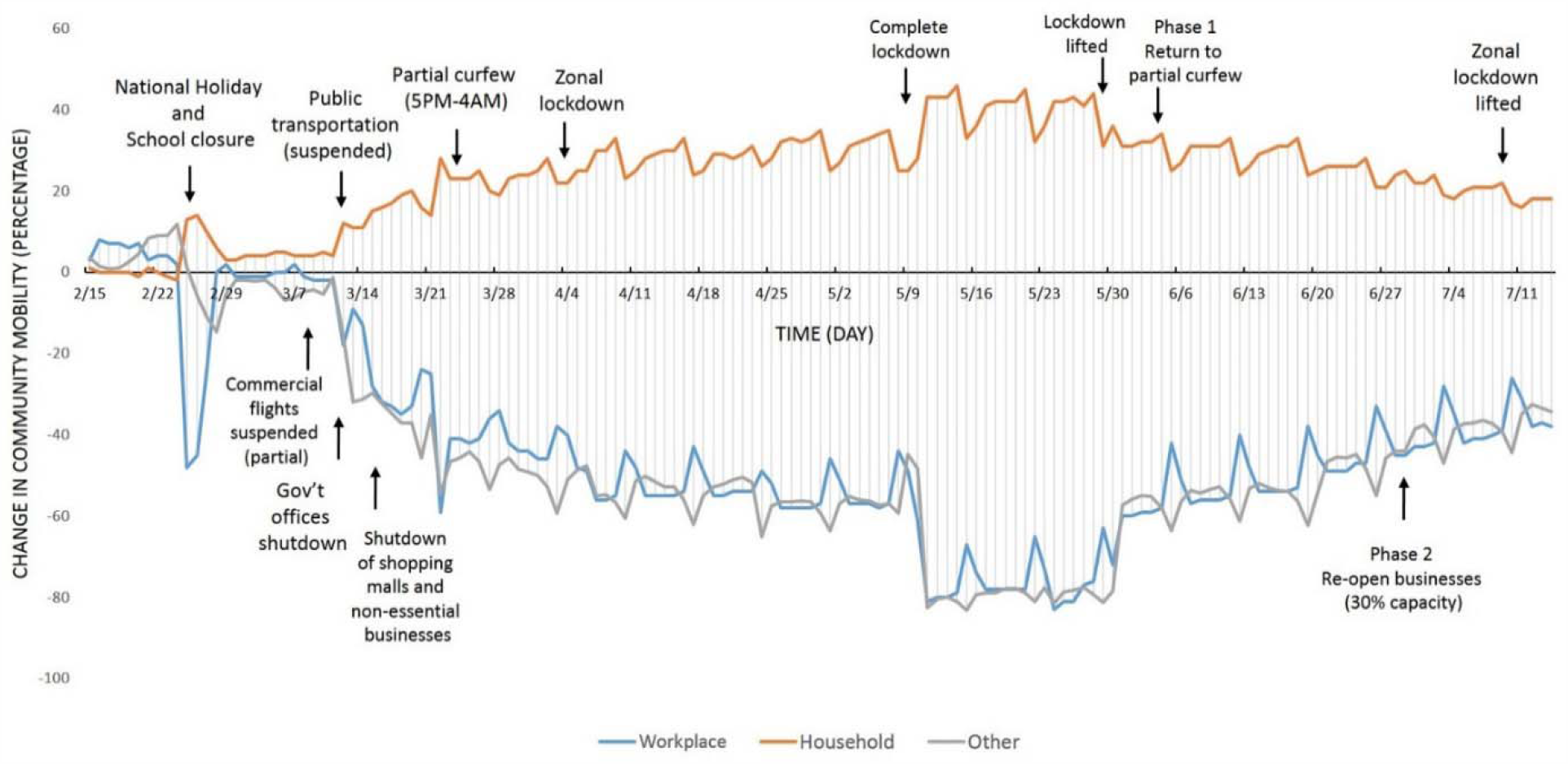
A timeline of Kuwaiti intervention measures and impacts to social mobility shown as percentage change in community mobility for workplace (blue line), household (orange line) and other (purple line) that affect social contacts. Google mobility data from February 15 to July 15 shown.

Based on estimation of the parameters of the model we have performed a scenario analyses and sensitivity analysis. If the lockdown was not activated and the mobility observed on day 76 and beyond continued then the expected epidemic peak would have happened on July 3, 2020.

### 3.2. Scenario Analysis

#### 3.2.1. Scenario 1

No school closures compared to school shutdowns if applied on day 1 of the outbreak (Feb. 24, 2020) (Figure 5).

**Figure 5.**
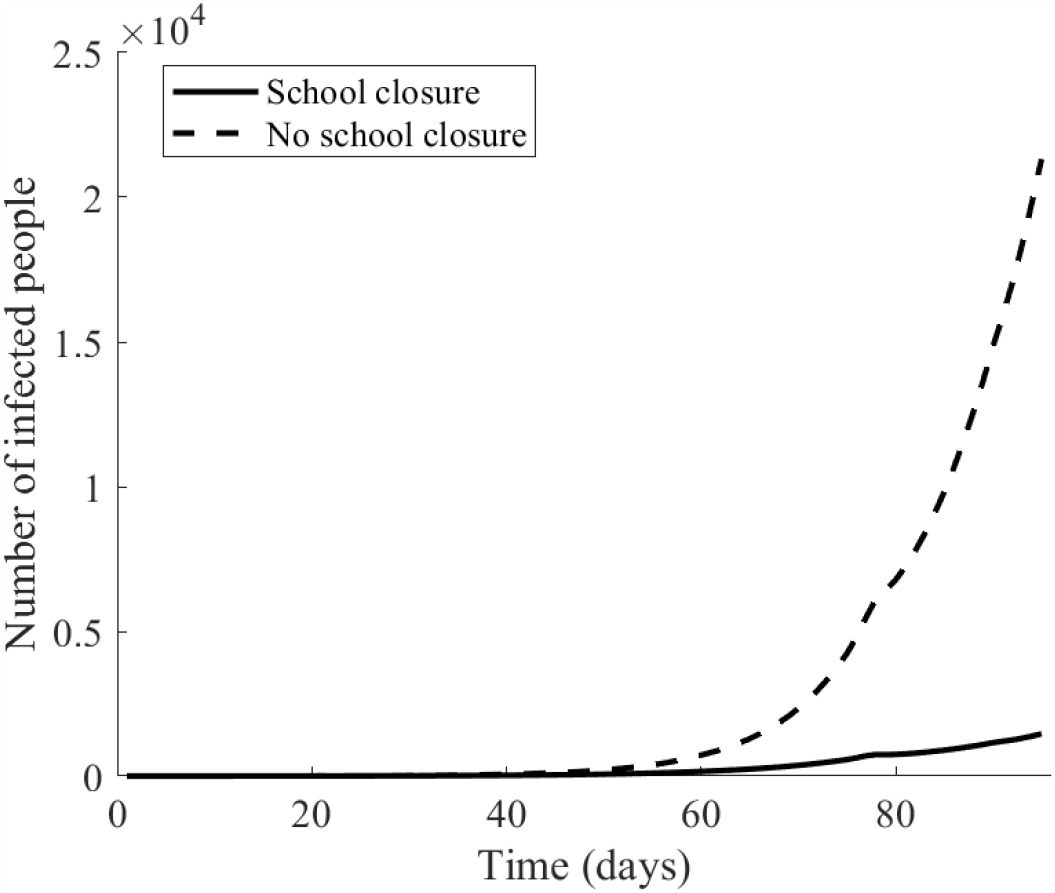
The number of infected people (non-cumulative cases) resulting from either school closures or no school closures. The start of the school closure began on day 8 (March 1, 2020) with the intervention in place until day 96, the end of the lockdown.

The comparison of school closures versus no school closures and its effect on the total cases from day 8 (March 1, 2020) until day 96 (May 31, 2020) shows that the school closure event resulted in a 5.55 multiple reduction in cases.

#### 3.2.2. Scenario 2

Shutdown of and non-essential services and government offices if applied on day 1 of the outbreak (Feb. 24, 2020) (Figure 6).

**Figure 6.**
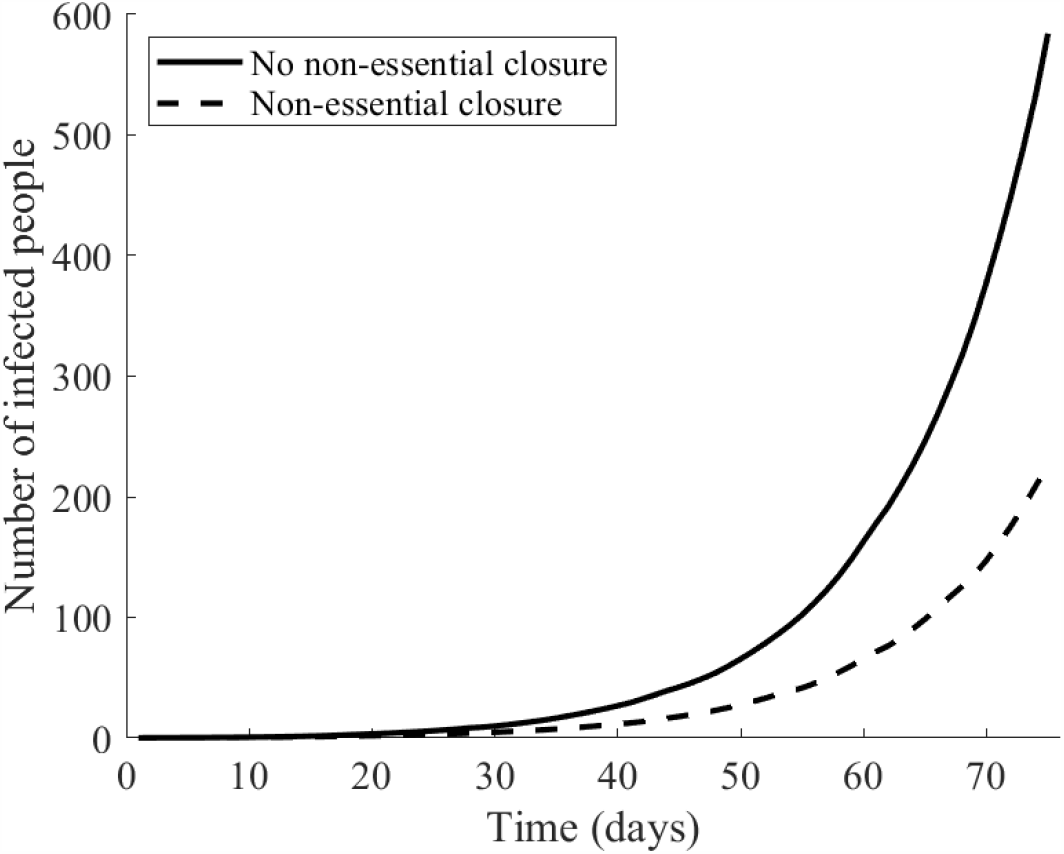
The number of infected people (non-cumulative cases) over time shown with and without closure of non-essential businesses, if these closures were implemented on day 1.

The government of Kuwait implemented a number of closures within the first three weeks of the COVID-19 outbreak including the closure of non-essential businesses (eg. cinemas, wedding halls, retail businesses, and eat-in restaurants) on March 1; shutdown of government offices on March 12; preventing mass gatherings (eg. prayers at the mosques) on March 13; and shutting shopping centers on March 15. The estimated contact reduction achieved by these movement restrictions was about 60% based on Google mobility data. The intervention to close non-essential services and businesses resulted in a 2.49 multiple drop in cases.

#### 3.2.3. Scenario 3

Lockdown versus no lockdown (Figure 7) and estimating the outbreak peak from fitted data and applying a lockdown under various timings and durations (Figure 8).

**Figure 7.**
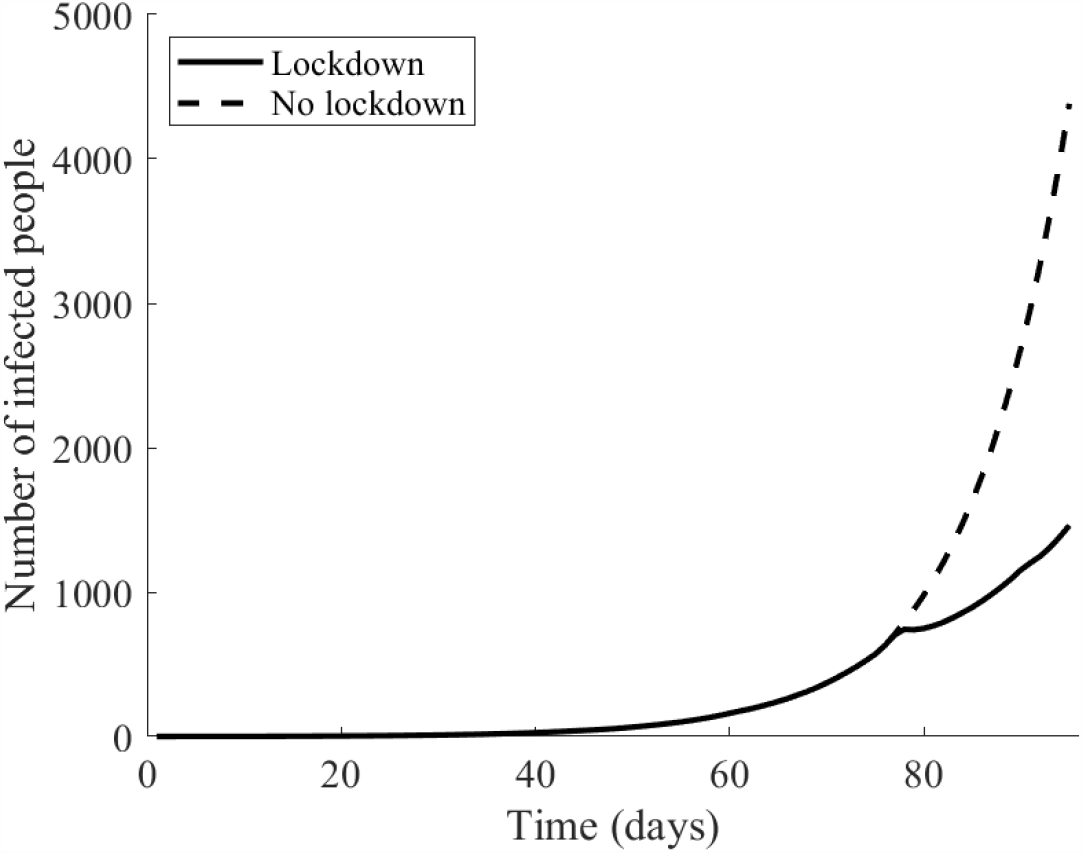
The number of non-cumulative cases (infected people) over time comparing the fitted data to a no-lockdown scenario starting on day 76 of the Kuwait COVID-19 outbreak.

**Figure 8.**
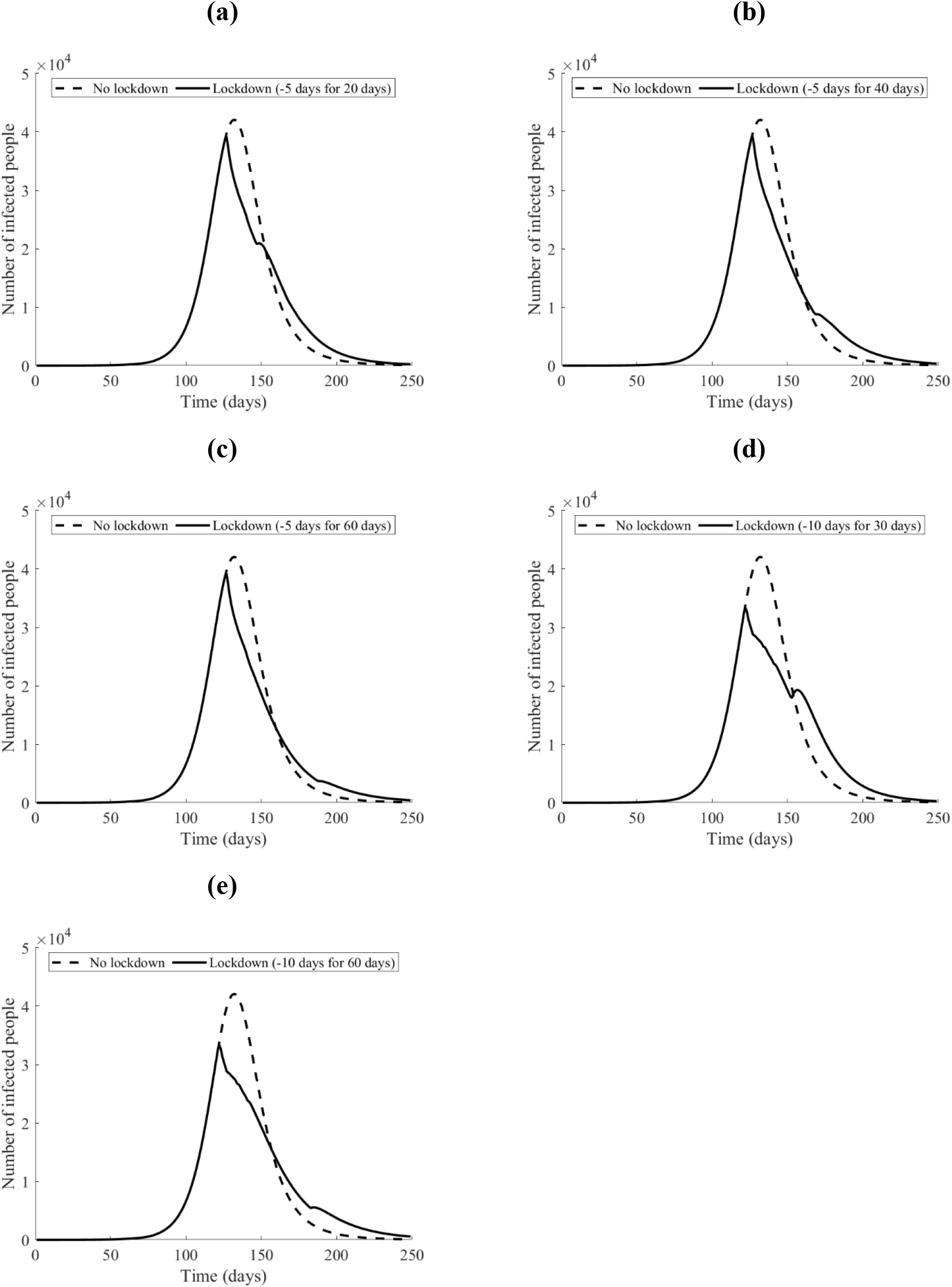
The number of infected people over time (non-cumulative, solid line) when full lockdowns are applied for (a) 20-days (b) 40-days and (c) 60-days beginning 5 days before the estimated peak or (d) 30-days and (e) 60-days of lockdown beginning 10 days before the estimated peak. In each case the lockdowns are compared to the number of people infected without a lockdown (non-cumulative, dashed line).

The government of Kuwait implemented a complete lockdown starting on May 10, 2020 (day 76) until May 31, 2020 (day 96). The effect of the full lockdown using Google mobility data showed increased home contact rates 20-35%, reduced work contact rate to 80%, reduced community contact rate by 80% as only grocery stores and essential services were allowed (Google, 2020). The action of the lockdown resulted in a 1.78 fold reduction in the number of cases.

Early modelling work predicted the peak of the first COVID-19 wave to occur in early May (Al-Shammari et al., 2020) and the government of Kuwait implemented its full lockdown to coincide with this estimate. With the availability of early case data our model effort with data fitting shows the COVID-19 peak was estimated to occur on day 132 (July 3, 2020). We show the effect of implementing the full lockdown for various durations (20, 40 or 60-days) starting either 5 days before the estimated peak (Figure 8, panels a-c) or 10 days before the estimated peak for 30 or 60-days duration (Figure 8, panels d-e).

We modeled the same 20-day lockdown (Figure 8, panel a) to reflect what realistically happened and we used the actual mobility data to model the movement and contact changes that occurred in Kuwait during the lockdown and in the days afterwards. The lockdown for 20 days if applied to the estimated peak resulted in a very modest 1.03 fold reduction. Longer durations up to 60 days (Figure 8, panel b-c) yielded similar results. Enacting the lockdown 10 days prior to the estimated peak for up either 30- or 60-days (Figure 8, panel d-e) while still showing modest fold peak reduction showed an improved attenuation of the curve with reduced non-cumulative case numbers.

#### 3.2.4. Scenario 4: Curfew versus no curfew (Figure 9)

The government of Kuwait implemented a partial curfew starting on March 22, 2020 and extended the curfew hours on April 7. A full curfew was put in place on May 10 to coincide with the start of the lockdown period for maximal contact reduction. The curfew hours were slowly relaxed over the next several months, through Phases 2-4, and finally the curfew was ended on August 30, 2020. We modelled the effect of a curfew versus no curfew scenario to show its effects if applied over the first 96 days.

**Figure 9.**
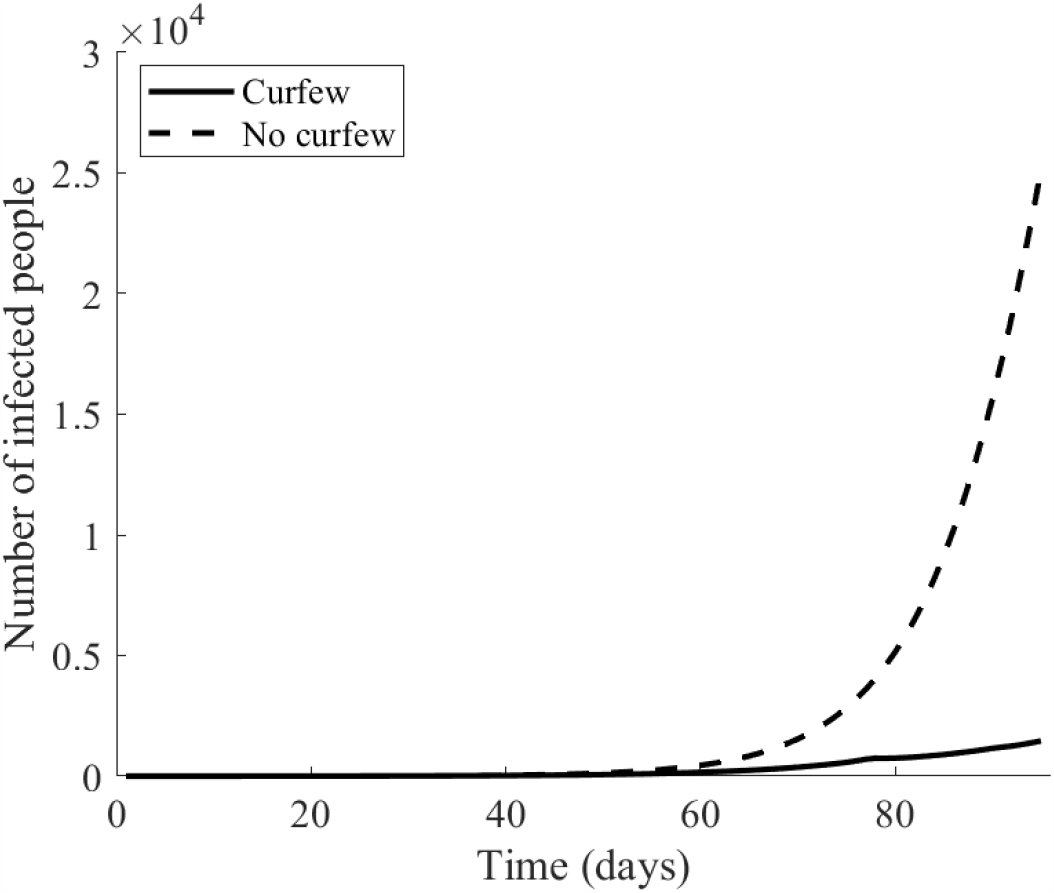
The number of infected people (non-cumulative cases) for either curfew or no curfew from March 22, 2020 until May 31, 2020 (day 96, the end of the lockdown intervention).

## 4. Discussion

We considered various scenarios with and without interventions to determine the impact of the interventions in reducing the numbers of cases.

### 4.1. Scenario 1: Impact of school closures

Our scenario compares no school closures to school shutdowns if applied on day 8 of the outbreak (March 1, 2020). The incidence of COVID-19 is far less in children than in adults. Nevertheless there are concerns about asymptomatic or mild paediatric cases going undetected and unknowingly transmitting SARS-CoV-2 in the community, inter-generationally and in schools to teachers, staff and other students (Qui et al., 2020). Additionally, the elderly in Kuwait’s families typically live together with younger generations in one dwelling, increasing the concern of transmission from asymptomatic or mildly symptomatic children to their elderly relatives. The epidemic dynamics of school closures and re-openings remains difficult to assess. A study that used social-contact data of school-aged children in an individual-based stochastic model in the USA found that school closures prevented a similar number of infections as workplace closures and social distancing measures by adults. The stochastic model estimated that school re-openings could increase symptomatic illness among middle and high school teachers of approximately 40% and about 4% among elementary school teachers. The transmission from children to other age groups was highly dependent on parameters with large uncertainty such as the relative susceptibility and infectiousness of children, and the extent of community transmissions occurring at the time of the school re-opening.

The first NPI used in Kuwait was the closure of schools and universities on March 1, 2020. Our model confirms the work by Head et al., (2020) with early school closures that had a positive effect of reducing COVID-19 transmission by 5.55 fold.

Decision-makers can implement a number of interventions upon school re-openings including reduced class sizes (cohorts of 20 elementary school students and cohorts of 10 middle or high school students) (Head et al., 2020) and full time and a part-time rotational class strategies with 50% of students attending school with at-home learning on alternate days or weeks as a way to reduce student density in the classrooms (Panovska-Griffiths et al., 2020).

Increased testing and contact tracing of both students and teachers, increased environmental cleaning, socially-distanced desk arrangements, distance learning, masking wearing while away from desks and maintaining social distance are some of the multiple in-school interventions strategies that also can be used when schools re-open (Johansen et al., 2020; Simon et al., 2020; Zhang et al., 2020).

### 4.2. Scenario 2: Impact of non-essential business closures

Kuwait closed non-essential services to slow down the distribution of COVID-19 in urban areas. The shutdown of non-essential services occurred by the third week of the epidemic with the closing of cinemas, wedding halls, and non-essential business venues (March 1, 2020); government office closings (March 12, 2020) and shutting down shopping centres (March 15, 2020). The reduction in social conduct resulting from non-essential business closures resulted in an estimated 2.49 fold reduction in cases.

In Wuhan, China the use of physical distancing with a staggered return to work after business closures was calculated to be one of the most effective strategies for non-essential business re-opening, with a projected reduction of the median number of infections by 92% (Prem et al., 2020). This was similar to Kuwait’s highly effective, gradual non-essential business re-opening strategy with an initial 30% return to work (Phase II, starting June 30, 2020); then hotel and mosque re-opening (Phase III, starting July 28, 2020); and government and private sectors re-opened with 50% capacity of their workforce (Phase IV, starting August 18, 2020).

### 4.3. Scenario 3: Lockdown versus no lockdown

A country-level analysis measuring the impact of government actions showed that full lockdowns when compared to partial lockdowns found that full lockdowns were strongly associated with recovery rates (as measured by recovered cases per million people) (Chaudhry et al, 2020). However, the timing and duration of the lockdown are critical factors to realize the benefits of this intervention.

Previously we showed hypothetical modeling of a lockdown in Kuwait timed 5-10 days before the estimated peak for 90-days in length yielded the optimal reduction in actual incidence and hospitalization. (Al-Zoughool et al., 2020; Oraby et al., 2020). Such lengthy lockdowns, while optimal, may not be practical resulting in devastating economic and psychosocial impacts. According to our previous hypothetical stochastic modelling for Kuwait (Al-Zoughool et al., 2020) a 20-day lockdown even if timed appropriately was likely too short to have much effect in reducing the first peak. Our modelled scenario (Figure 8 panel a) considered a similar 20-day lockdown beginning 5-days prior to the peak which we estimated to be on July 3, 2020. We did not observe a major reduction in cases or a “tunneling effect” shown by a reduced number of cases that would be split or bypass the peak from a 20-day lockdown. Previous hypothetical modelling showed that optimal results and a tunneling effect could be achieved with a 90-day lockdown (Al-Zoughool et al., 2020), while shorter duration lockdowns of 40-45 days, if timed accurately, could also realize significant impacts to reducing case numbers and hospitalizations. Our results suggest the 20-day lockdown implemented too far ahead of the actual peak was likely only minimally effective.

We investigated longer lockdown durations of 40-days and 60-days starting 5 days before the peak (Figure 8, panel b and c) which also appeared to have limited effects. However, 30-days and 60-days of lockdown starting 10 days before the peak (Figure 8, panel d-e) both showed a tunneling effect and a 25% reduction in the overall number of cases. Thus, a well-timed lockdown initiated 10 days before the estimated peak for 30 to 60 days would bypass the peak. This result is significant as it has the effect of reducing overall numbers of infections and subsequently the numbers of severe cases entering hospitals which would help prevent exceeding ICU bed availability and overwhelming available hospital resources.

### 4.4. Scenario 4: Curfew compared to no curfew

Curfew is a government mandated stay-at-home order limited to specific hours of the day as a way to decrease contact between people and reduce community transmission of COVID-19. Comparative analysis of curfews in Jordan compared to Kuwait and other gulf countries showed using a country-wide curfew can be an effective NPI to reduce the spread of COVID-19 – if implemented early with country-wide compliance (Khatatbeh, 2020). Kuwait enacted its first curfew on March 22, 2020 (28 days after the country’s first confirmed case), by this time 176 confirmed cases already had been reported (Worldometer, 2020).

Comparing the intervention effects of a curfew versus no curfew showed that curfews can be highly effective and similar to school closures when applied early. Figure 9 showed that limiting the movement of people through curfews resulted in an estimated 6.60-fold reduction in cases.

## 5. Conclusion

The World Health Organization (WHO) officially declared the coronavirus outbreak a pandemic on March 11, 2020 (Cucinotta and Vanelli, 2020). Several countries, including Kuwait, had already quickly mobilized to respond to the emerging outbreak ahead of the WHO announcement. Of the NPIs we reviewed (school closures, non-essential business closures, curfews, and lockdown) many were implemented by almost all countries to some degree by the end of March 2020 (Gollwitzer et al., 2020).

The SEAIR model we developed to review these early interventions captures and reflects this first exponential phase of the COVID-19 epidemic in Kuwait when the system was homogeneous. We used modified social contact rates refined using Google mobility data to better reflect changes in social contact rates in Kuwait. The data fitting analysis allowed for calculation of the basic reproduction number *R*_0_(t). We found that the *R*_0_(t) was significantly dampened to about a third of its initial value. This affirms that the early intervention measures implemented had a dramatic (and substantial) effect in attenuating the COVID-19 outbreak in Kuwait.

The model also gives insights into what might have happened under different “what-if” scenarios which can inform future control policy and decision-making in the event of future waves. Until COVID-19 vaccines are more widely available and delivered to the population, the use of non-pharmaceutical initiatives (NPIs) such as contract tracing with case isolation, school closures, non-essential business closures, government office shut downs, banning mass gatherings and public events, curfews and lockdowns are the best available options to restrict the spread of SARS-CoV-2.

Kuwait implemented a series of NPIs within the first few weeks of confirmed cases entering the country and early use of interventions such as school closures, non-essential business closures and curfews were highly effective in reducing case numbers. Our model suggests that, in retrospect, the three-week lockdown would likely have been even more effective in alleviating later hospital caseloads if its implementation were delayed to a time closer to the epidemic peak. Such considerations, however, were impossible to know or predict at that time. However, these prediction model results, based on real data from Kuwait, can help policy-makers in making lockdown decisions in future outbreaks of COVID-19 or other infections.

All of the intervention measures aim at reducing the rate of infection transmission in the community which both delays and reduces the magnitude of the epidemic peak. The actions serve two purposes, first to allow additional time for the healthcare system to prepare and respond efficiently to the pandemic wave and second, to manage the outbreak until the development and deployment of potential new treatments and vaccines.

Finally, while the fitted case data provides a good retrospective review of the effects of NPIs for the first wave of COVID-19 in Kuwait the lessons learned can be carried forward and applied to future pandemic waves. Some NPIs (restrictions that create social distancing outside the household or reduce movement of people) when applied early can be highly effective while other interventions (full lockdown) appear to require more precision in their timing and duration.

## Data Availability

Contact Dr. Mustafa Al-Zoughool regarding the availability of data referred to in the manuscript

## 6. Acknowledgement

Dr. Tamer Oraby dedicates this work to the soul of Dr. Mokhtar Konsowa (1953-2012) whose mentorship and kindness will never be forgotten.

## 7. Author’s contributions

MT: participated in the study design, helped parameterize the model and helped draft the manuscript and critically revised it.

TO: participated in the design of the study and the analyses plan, conducted the modeling and coded it, carried out the data analyses, helped draft the manuscript, and critically revised the manuscript;

JL: contributed to the study design, collected and collated case data, and reviewed the manuscript

HV: contributed to the study design and critically revised the manuscript.

JG: contributed to the study design and critically revised the manuscript.

WAA: contributed to the design of the study, the analyses plan and manuscript review.

MAA: contributed to the study design, reviewed the analyses, results and reviewed the manuscript

SE: contributed to the study design, helped parameterize the model, and critically revised the manuscript.

MAZ: Conceived the study, participated in the design of the study, helped parameterize the model, coordinated the study, guided the analysis plan, reviewed the results, and helped both write and critically review the manuscript.

All authors gave final approval for publication.

## Acknowledgement and source of funding

This study was supported by Kuwait Institute for Advancement of Science (KFAS) grant number CORONA-46 to Dr. Al-Zoughool.

## Notes

### Competing Interest Statement

The authors have declared no competing interest.

